# Association of hypotension with mortality among US adults: prospective cohort study

**DOI:** 10.1101/2024.11.08.24316952

**Authors:** Yan Han, Jing Tang, Na Wu, Zhao Li, Dachuan Cai, Hong Ren, Peng Hu, Zhiwei Chen

**Affiliations:** Department of Infectious Diseases, the Second Affiliated Hospital of Chongqing Medical University, Chongqing, China; Key Laboratory of Molecular Biology for Infectious Diseases (Ministry of Education), Institute for Viral Hepatitis, Chongqing Medical University, Chongqing, China; Department of Gastroenterology, the Seventh People’s Hospital of Chongqing, Chongqing, China; Department of Infectious Diseases, the First Affiliated Hospital of Chongqing Medical University, Chongqing, China

**Author notes:** Corresponding authors: Zhiwei Chen, Institute for Viral Hepatitis, The Key Laboratory of Molecular Biology for Infectious Diseases, Chinese Ministry of Education, The Second Affiliated Hospital of Chongqing Medical University, 74 Linjiang Road, Yuzhong District, Chongqing 400010, China. E-mail addresses; Peng Hu, Key Laboratory of Molecular Biology for Infectious Diseases (Ministry of Education), Institute for Viral Hepatitis, Chongqing Medical University, Chongqing, China; Department of Infectious Diseases, the First Affiliated Hospital of Chongqing Medical University, Chongqing, China, 1 Youyi Road, Jiulongpo District, Chongqing 400016, China. E-mail addresses. Hong Ren, Institute for Viral Hepatitis, The Key Laboratory of Molecular Biology for Infectious Diseases, Chinese Ministry of Education, The Second Affiliated Hospital of Chongqing Medical University, 74 Linjiang Road, Yuzhong District, Chongqing 400010, China. E-mail addresses. Joint first authors: contributed equally (Yan Han, Jing Tang).

## Abstract

**Background:** The associations of hypotension with mortality in general population remains incompletely understood. We aimed to investigate whether hypotension is associated with higher all-cause and cardiovascular disease (CVD) mortality in this population.

**Methods:** In this prospective analysis, we utilized data from the National Health and Nutrition Examination Survey (NHANES, 1999-2018), with mortality information linked until 2019. We used multivariable Cox proportional hazards regression to estimate hazard ratios (HRs) and 95% CIs for the associations of different blood pressure (BP) with all-cause and CVD mortality.

**Findings:** Among the 37,832 participants, a total of 5261 deaths and 1664 deaths attributed to CVD causes were recorded over a median of 8.4 years of follow-up. The prevalence of hypotension was 7.6%. Both systolic BP and diastolic BP exhibited a J-shaped association with the all-cause and CVD mortality in restricted cubic spline modeling analysis (nonlinear-P <0.01). Compared to the normal BP group, the adjusted HRs for all-cause and CVD mortality in the hypotension group were 1.44 (1.20-1.74) and 1.57 (1.10-2.24), respectively. Subgroup analyses revealed that older individuals (age ≥60 years) and those with obesity exhibited more pronounced HRs for all-cause mortality, with HRs of 1.60 (1.28-2.00) and 1.95 (1.45-2.61), respectively (P for interaction <0.05).

**Interpretation:** In this nationally representative cohort of US adults, hypotension demonstrated a significant association with both all-cause and cardiovascular disease mortality, particularly among elderly and obesity individuals. The findings underscore the significance of paying attention to and optimizing the management of hypotension in the general population.

## Introduction

Benefiting from extensive research and education, the global awareness of hypertension has significantly increased.^1–4^ However, hypotension, a relatively benign condition, often goes unnoticed mainly because it is typically asymptomatic.^5^ Several studies conducted in the early 1980s yielded disparate findings regarding the association between mortality and low blood pressure (BP).^6,7^ Moreover, a study involving individuals aged 65 years and older in Norway found no significant correlation between low systolic BP (SBP) and increased mortality,^8^ however, the Leiden 85-plus study reported that low SBP predicts an elevated risk of mortality among subjects aged 90 years.^9^ Consequently, there remains controversy surrounding whether hypotension increases mortality risk within the general population. In recent years, research on hypotension has primarily focused on specific populations such as sepsis patients, critically ill individuals, and those undergoing hemodialysis.^10–13^ Nevertheless, the relationship between hypotension and mortality in the general population still lacks clarity.

In this national prospective observational cohort study, we utilized data from 1999-2018 cycles of the National Health and Nutrition Examination Survey (NHANES) to investigate the association between hypotension and all-cause as well as cardiovascular disease (CVD) mortality among adults in the United States (US).

## Methods

### Study population

The NHANES is a series of cross-sectional surveys conducted by the National Center for Health Statistics (NCHS) using a complex, multistage probability sample design to collect health-related information about the civilian noninstitutionalized population in the US.^14^ The NHANES was authorized by the NCHS ethics review board, and written informed consent was obtained from all participants.^15^ This study adhered to the Strengthening the Reporting of Observational Studies in Epidemiology (STROBE) reporting guideline.

We utilized data from ten consecutive cycles of the NHANES spanning from 1999 to 2018. Among a total of 55,081 adults aged ≥20 years, we excluded individuals who met the following criteria: (1) missing data of BP (n = 5,109), (2) incomplete data on potential covariates (n = 11,266), and (3) pregnant individuals (n = 825), and (4) those with unclear vital status due to being ineligible for mortality record linkage (n = 49). Consequently, our final analytical sample consisted of 37,832 participants included in this study (eFigure 1 in Supplement 1). Minor discrepancies were observed in several baseline characteristics such as age and sex when comparing the included participants with the entire population (eTable 1 in Supplement 1).

### Classification of blood pressure

All BP determinations (systolic and diastolic) were conducted at the mobile examination center of NHANES. Following a period of quiet rest in a seated position for five minutes, three consecutive BP readings were obtained. According to the European Society of Cardiology (ESC) definition,^16^ hypertension was defined as an average SBP ≥140 mmHg and/or diastolic BP (DBP) ≥90 mmHg, or the use of antihypertensive drugs. High normal BP was defined as 130-139 and/or 85-89 mmHg. As there is no standard definition for hypotension, we considered values below <90 or <60 mmHg as indicative of hypotension in this study.^5^ Accordingly, normal BP was defined as 90-129 and/or 60-84 mmHg.

### Outcomes and covariates

The primary outcome of the study was mortality due to all causes and CVD, which was assessed using the National Death Index until December 31, 2019.^17^ Deaths with codes in the range I00–I99 were classified as originating from cardiovascular conditions. Time to event (in months) was calculated from the NHANES interview day to either the death date or the end of follow-up (December 31, 2019), whichever occurred first.

Covariates in this study encompassed age, sex, race/ethnicity (Mexican-American, other Hispanic, non-Hispanic White, non-Hispanic Black, and other race), educational attainment (high school or less, college or associates degree, and college graduate or above), marital status (married and unmarried), smoking status (yes and no), drinking status (yes and no), physical activity (yes and no), history of CVD (yes and no), history of chronic lung diseases (yes and no), history of cancer (yes and no), diabetes mellitus (yes and no), hypercholesterolemia (yes and no), body mass index (BMI), alanine aminotransferase (ALT) levels, and estimated glomerular filtration rate (eGFR) levels. Detailed definitions of these covariates were provided in the eMethods in Supplement 1.

### Subgroup and sensitivity analyses

Subgroup analyses were conducted to further investigate the main findings, stratifying by age (<60 and ≥60 years), sex, race/ethnicity, diabetes mellitus, hypercholesterolemia, BMI (<25, 25-29.9, ≥30 kg/m^2^) and eGFR (<90 and ≥90 mL/min/1.73m^2^). Sensitivity analyses were also performed to assess result robustness. Firstly, we employed a more stringent definition of hypertension from the American College of Cardiology (ACC): SBP ≥130 mmHg or DBP ≥80 mmHg.^18^ High normal BP was defined as 120-129 and <80 mmHg, which companied with the change of normal BP, i.e., 90-119 and 60-79 mmHg. Secondly, regression analysis included the poverty income ratio and healthy eating index as covariates. Lastly, participants who experienced mortality within the first years of follow-up were excluded to minimize potential bias.

### Statistical analysis

Given the NHANES complex sampling method, all analyses in this study incorporated sample weights, clustering, and stratification. Categorical variables were reported as percentages, while continuous variables were presented as means accompanied by standard deviations (SDs). Comparisons of baseline characteristics between different BP groups were conducted using χ^2^ tests for categorical variables and t-tests for continuous variables. Moreover, restricted cubic spline models with 5 knots (at the 5th, 27.5th, 50th, 72.5th, and 95th percentiles) were fitted to explore potential nonlinear associations of continuous SBP and DBP values with all-cause and CVD mortality outcomes. The likelihood ratio test was performed for the nonlinearity testing.

The hazard ratios (HRs) and 95% confidence intervals (CIs) for all-cause and CVD mortality associated with BP were estimated using multivariable Cox proportional hazards regression models. Model 1 was adjusted for age, sex, race/ethnicity, education level, marital status, smoking status, drinking status, and physical activity. Model 2 was additionally adjusted for history of CVD, history of chronic lung diseases, history of cancer, diabetes mellitus, hypercholesterolemia, BMI, alanine aminotransferase, and eGFR levels. Additionally, heat maps were constructed to visually represent the impact of different combinations of BP on their associations with outcomes.

The statistical analysis was performed using R software, version 4.3.2 (R Foundation for Statistical Computing). All hypothesis tests were two-tailed, and a significance level of P < 0.05 was employed.

## Results

### Participant Characteristics

As presented in Table 1, the analytic sample consisted of 37,832 participants (mean [SD] age, 47.58 [16.88] years; 19,070 men [49.6%]). The prevalence rates for hypertension, high normal BP, normal BP, and hypotension was 38.2%, 9.3%, 44.9%, and 7.6%, respectively. In comparison to individuals with normal BP, those with hypotension exhibited younger, women, lower proportions of diabetes mellitus and hypercholesterolemia, and higher eGFR levels. Conversely, contrasting characteristics were observed among individuals with hypertension. (Table 1)

**Table 1.** Characteristics of participants in the National Health and Nutrition Examination Survey, 1999-2018.

| **Characteristic** | **Overall**<br>**(N=37,832)** | **Hypotension**<br>**(N=2,856)** | **Normal BP**<br>**(N=16,978)** | **High Normal BP**<br>**(N=3,536)** | **Hypertension**<br>**(N=14,462)** | **P value** |
| --- | --- | --- | --- | --- | --- | --- |
| Age, years | 47.58 (16.88) | 37.55 (16.80) | 41.14 (14.10) | 48.85 (15.59) | 59.66 (14.06) | <0.001 |
| Men | 19,070 (49.6%) | 1,186 (41.0%) | 8,478 (49.2%) | 2,181 (60.7%) | 7,225 (49.2%) | <0.001 |
| Race and ethnicity |  |  |  |  |  | <0.001 |
| Mexican American | 6,492 (8.2%) | 602 (11.9%) | 3,231 (9.4%) | 647 (8.5%) | 2,012 (5.3%) |  |
| Other Hispanic | 3,279 (5.4%) | 309 (7.7%) | 1,584 (5.9%) | 285 (5.1%) | 1,101 (4.1%) |  |
| Non-Hispanic White | 16,998 (69.6%) | 1,217 (64.3%) | 7,564 (68.9%) | 1,566 (69.5%) | 6,651 (71.8%) |  |
| Non-Hispanic Black | 7,610 (10.2%) | 494 (10.1%) | 2,743 (8.4%) | 737 (11.0%) | 3,636 (13.0%) |  |
| Other race | 3,453 (6.6%) | 234 (6.0%) | 1,856 (7.3%) | 301 (6.0%) | 1,062 (5.8%) |  |
| Education level |  |  |  |  |  | <0.001 |
| High school or less | 18,583 (40.3%) | 1,369 (39.4%) | 7,455 (36.4%) | 1,811 (42.5%) | 7,948 (46.0%) |  |
| College or associates degree | 10,968 (31.4%) | 882 (32.8%) | 5,128 (31.3%) | 1,015 (32.5%) | 3,943 (30.8%) |  |
| College graduate or above | 8,281 (28.3%) | 605 (27.8%) | 4,395 (32.2%) | 710 (25.0%) | 2,571 (23.1%) |  |
| Married | 22,729 (63.9%) | 1,566 (56.5%) | 10,472 (64.7%) | 2,176 (64.9%) | 8,515 (64.3%) | <0.001 |
| Smoker | 17,497 (46.5%) | 1,271 (46.3%) | 7,327 (43.9%) | 1,708 (48.3%) | 7,191 (50.1%) | <0.001 |
| Drinker | 29,172 (81.6%) | 2,290 (83.0%) | 13,724 (84.5%) | 2,781 (82.9%) | 10,377 (76.2%) | <0.001 |
| Sedentary | 9,294 (19.6%) | 585 (16.0%) | 3,243 (15.2%) | 797 (18.5%) | 4,669 (27.5%) | <0.001 |
| Diabetes mellitus | 6,523 (12.6%) | 212 (5.1%) | 1,404 (5.9%) | 495 (10.6%) | 4,412 (25.4%) | <0.001 |
| Hypercholesterolemia | 15,338 (39.4%) | 628 (22.4%) | 4,885 (29.3%) | 1,374 (39.9%) | 8,451 (59.1%) | <0.001 |
| History of cardiovascular disease | 4,249 (8.7%) | 193 (6.0%) | 689 (3.4%) | 260 (5.6%) | 3,107 (18.6%) | <0.001 |
| History of chronic lung disease | 2,768 (7.3%) | 185 (6.2%) | 919 (5.7%) | 234 (6.6%) | 1,430 (10.1%) | <0.001 |
| History of cancer | 3,590 (9.8%) | 168 (6.1%) | 978 (6.4%) | 312 (9.2%) | 2,132 (16.1%) | <0.001 |
| Body mass index, kg/m<sup>2</sup> | 28.87 (6.70) | 26.22 (5.49) | 27.78 (6.13) | 29.87 (6.94) | 30.91 (7.14) | <0.001 |
| eGFR, mL/min/1.73m<sup>2</sup> | 92.13 (21.38) | 101.59 (20.48) | 98.29 (18.33) | 92.44 (18.79) | 80.17 (21.48) | <0.001 |
| Alanine aminotransferase, U/L | 25.59 (20.38) | 22.09 (15.64) | 25.12 (20.17) | 28.83 (24.13) | 26.21 (20.36) | <0.001 |
| Systolic BP, mmHg | 122.76 (17.53) | 105.59 (8.75) | 114.42 (8.68) | 132.09 (5.36) | 137.21 (19.81) | <0.001 |
| Diastolic BP, mmHg | 70.96 (12.21) | 54.03 (6.75) | 70.65 (7.20) | 76.03 (11.45) | 74.02 (15.68) | <0.001 |

Note: Data are shown as the mean (SD) or unweighted frequency counts (weighted percentage) as appropriate. Abbreviations: BP, blood pressure; eGFR, estimated glomerular filtration rate.

### All-cause and cardiovascular disease mortality associated with continuous BP values

During a median follow-up period of 8.4 years (interquartile range, 4.6-12.4 years), a total of 5261 deaths were recorded, with 1664 deaths attributed to cardiovascular causes. Restricted cubic spline modeling revealed a J-shaped association between both SBP and DBP with all-cause mortality (Figure 1A and B). Two cutoff points for SBP were identified at 103 mmHg and 122 mmHg where the HR values were 1. This indicates that both SBP >122 mmHg or <103 mmHg were associated with an increased risk of all-cause mortality. Additionally, the lowest HR was observed at an SBP of 113 mmHg. For DBP, the two cutoff points were found at 70 and 83 mmHg respectively, while the breakpoint corresponding to the lowest HR was determined as 77 mmHg. Similar trends were observed in the relationship between BP and CVD mortality (Figure 1C and D).

**Figure 1.**
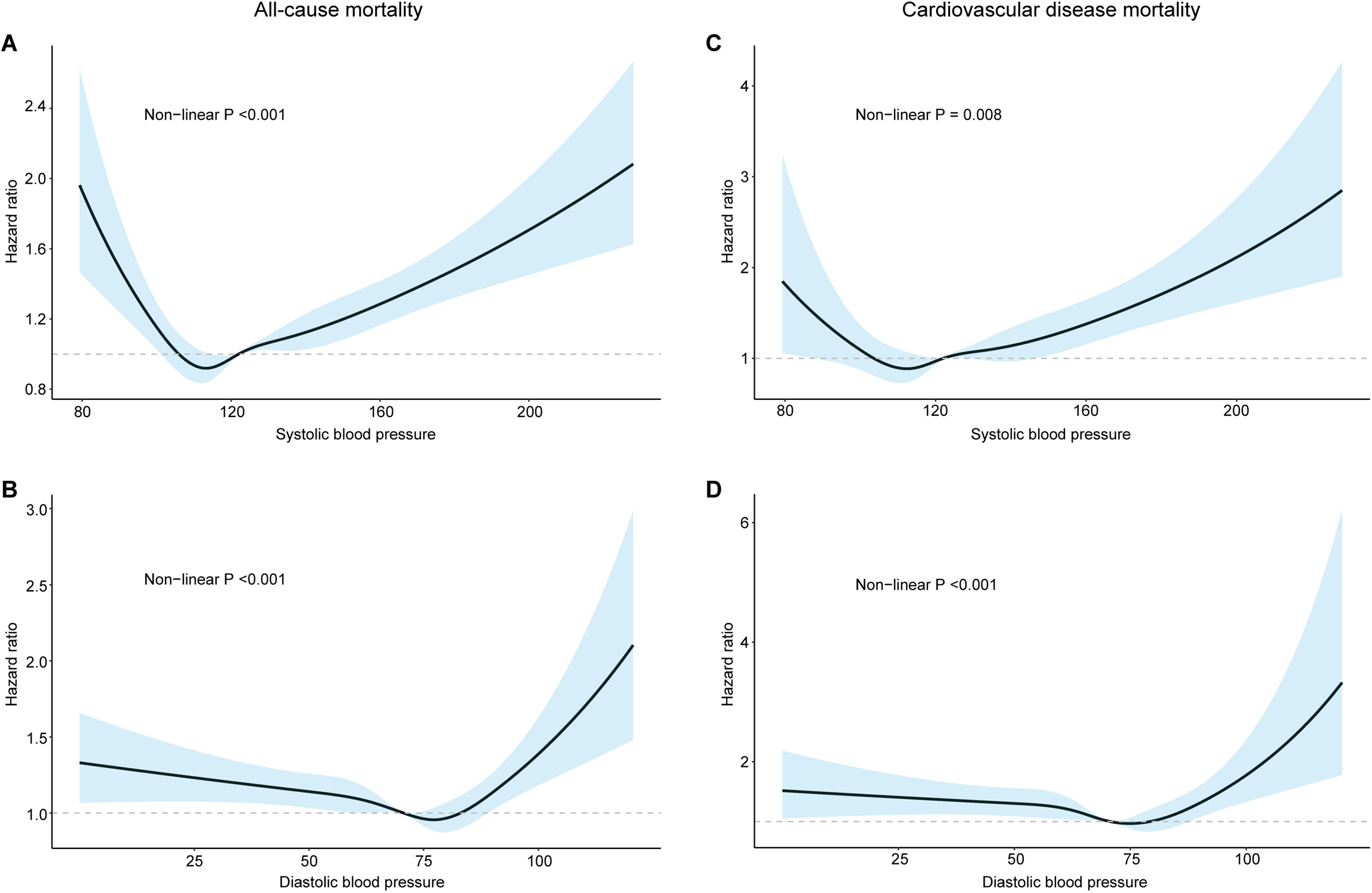
Association of BP as continuous variable with all-cause and CVD mortality using restricted cubic spline models. **(A)** Systolic BP and all-cause mortality. **(B)** Diastolic BP and all-cause mortality. **(C)** Systolic BP and CVD mortality. **(D)** Diastolic BP and CVD mortality. Hazard ratios (solid lines) and 95% confidence intervals (shaded areas) were estimated after adjusting for age, sex, race/ethnicity, education level, marital status, smoking status, drinking status, physical activity, history of CVD, history of chronic lung diseases, history of cancer, diabetes mellitus, hypercholesterolemia, body mass index, alanine aminotransferase, and estimated glomerular filtration rate levels. The restricted cubic spline regression models were conducted with 5 knots at the 5th, 27.5th, 50th, 72.5th, and 95th percentiles of systolic and diastolic BP. Abbreviations: BP, blood pressure; CVD, cardiovascular disease.

Interestingly, upon categorizing participants into low and high BP groups based on their breakpoints at the lowest HR, we observed that individuals in the low SBP group exhibited a decreased risk of all-cause mortality, with an HR of 0.74 (95% CI, 0.64-0.87) per 10 mmHg increment in BP after multivariable adjustment in model 2. Conversely, among those in the high SBP group, there was an inverse trend observed (HR, 1.08; 95% CI, 1.06-1.10). Similar findings were also noted for DBP groups.

Regarding CVD mortality, the trends aligned with those observed for all-cause mortality except for the low SBP group where statistical significance was not achieved. (eTable 2 in Supplement 1)

### All-cause and cardiovascular disease mortality associated with hypotension

When stratifying participants based on their combined SBP and DBP into different groups, we observed an increased HR for all-cause and CVD mortality among individuals with BP levels lower or higher than the reference range of 100-120/70-80 mmHg (Figure 2). Moreover, a greater deviation from the reference range was associated with a larger difference in HR. Additionally, BP was categorized into four groups according to predefined definitions. Notably, compared to the normal BP group, both hypertension and hypotension groups exhibited adjusted HRs of 1.32 (95% CI, 1.20-1.45) and 1.44 (95% CI, 1.20-1.74), respectively, for all-cause mortality (Table 2). Similarly, elevated adjusted HRs were also observed in both hypertension and hypotension groups compared to the normal BP group for CVD mortality (1.58 [95% CI, 1.28-1.95] and 1.57 [95% CI, 1.10-2.24], respectively; Table 2).

**Figure 2.**
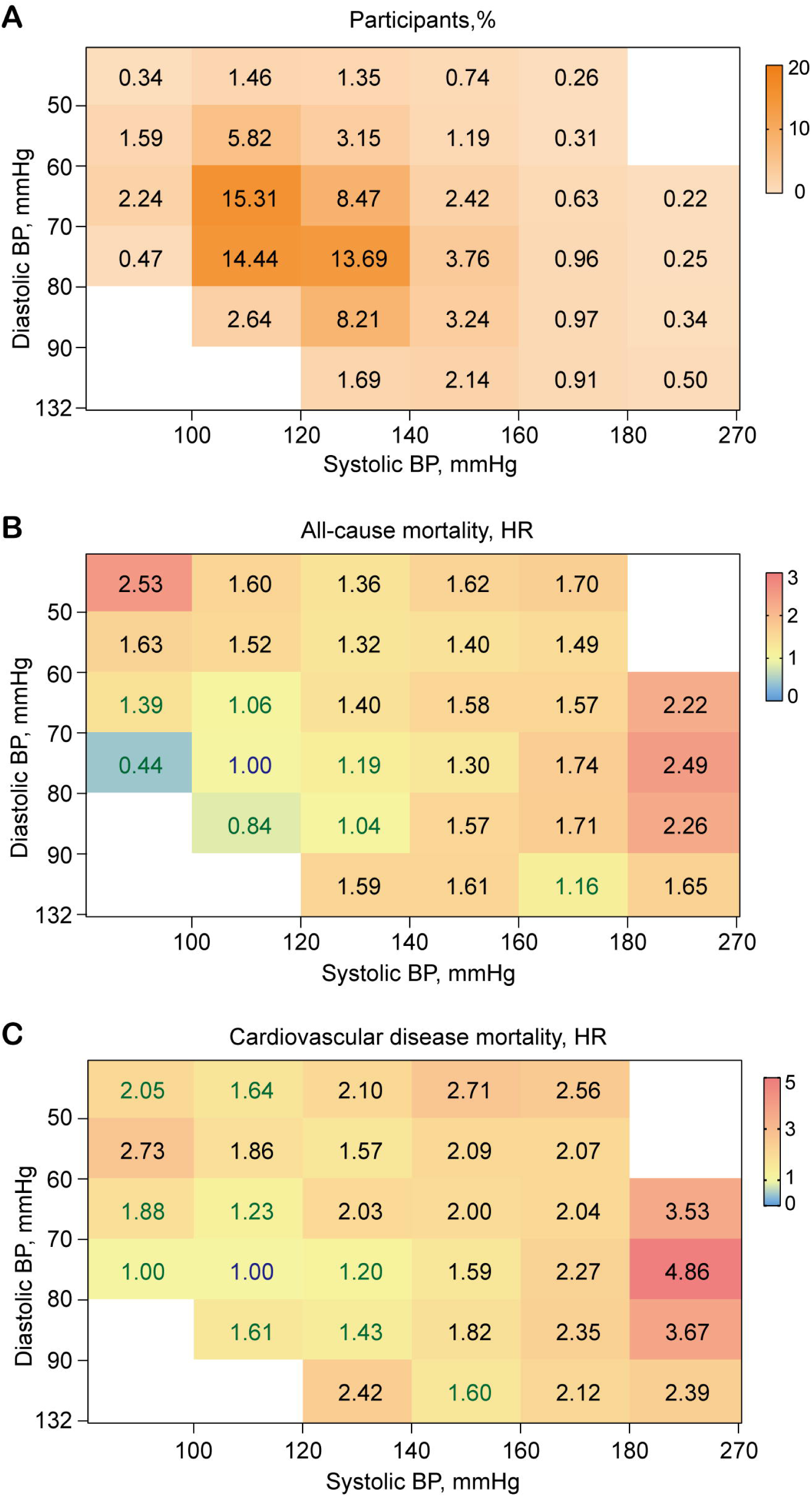
Heat maps depicting all-cause and CVD mortality in relation to different systolic and diastolic BP combinations. **(A)** Participants distribution. **(B)** HR distribution for all-cause mortality. **(C)** HR distribution for CVD mortality. Derived by Cox proportional hazards regression with systolic and diastolic BP analyzed as categorical variables. The reference group was defined as individuals with BP within the range of 100-120/70-80 mmHg. In panel A, the numbers represent the percentage of participants in each BP category. Groups with fewer than 50 actual numbers were excluded from both display and analysis. In other panels, the numbers represent HRs associated with the risk of mortality; blue numbers indicate the reference group, green numbers indicate non-significant HRs, and black numbers indicate statistically significant HRs. Abbreviations: BP, blood pressure; CVD, cardiovascular disease; HR, hazard ratio.

**Table 2.**
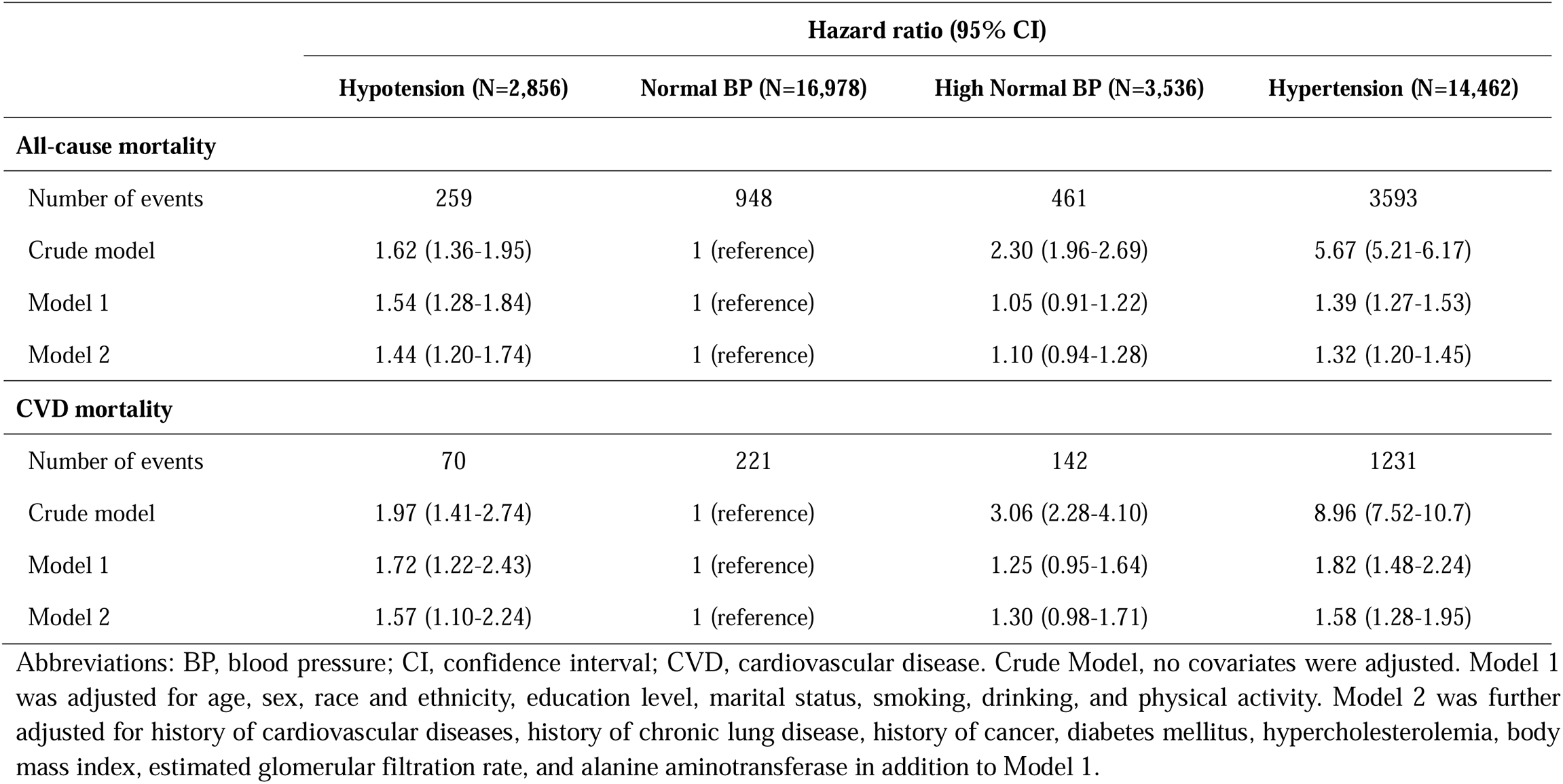
Associations of all-cause and CVD mortality with different classifications of blood pressure.

|  | Hazard ratio (95% CI) |  |  |  |
| --- | --- | --- | --- | --- |
|  | Hypotension (N=2,856) | Normal BP (N=16,978) | High Normal BP (N=3,536) | Hypertension (N=14,462) |
| **All-cause mortality** |  |  |  |  |
| Number of events | 259 | 948 | 461 | 3593 |
| Crude model | 1.62 (1.36-1.95) | 1 (reference) | 2.30 (1.96-2.69) | 5.67 (5.21-6.17) |
| Model 1 | 1.54 (1.28-1.84) | 1 (reference) | 1.05 (0.91-1.22) | 1.39 (1.27-1.53) |
| Model 2 | 1.44 (1.20-1.74) | 1 (reference) | 1.10 (0.94-1.28) | 1.32 (1.20-1.45) |
| **CVD mortality** |  |  |  |  |
| Number of events | 70 | 221 | 142 | 1231 |
| Crude model | 1.97 (1.41-2.74) | 1 (reference) | 3.06 (2.28-4.10) | 8.96 (7.52-10.7) |
| Model 1 | 1.72 (1.22-2.43) | 1 (reference) | 1.25 (0.95-1.64) | 1.82 (1.48-2.24) |
| Model 2 | 1.57 (1.10-2.24) | 1 (reference) | 1.30 (0.98-1.71) | 1.58 (1.28-1.95) |

Abbreviations: BP, blood pressure; CI, confidence interval; CVD, cardiovascular disease. Crude Model, no covariates were adjusted. Model 1 was adjusted for age, sex, race and ethnicity, education level, marital status, smoking, drinking, and physical activity. Model 2 was further adjusted for history of cardiovascular diseases, history of chronic lung disease, history of cancer, diabetes mellitus, hypercholesterolemia, body mass index, estimated glomerular filtration rate, and alanine aminotransferase in addition to Model 1.

### Subgroup Analyses

Subgroup analyses consistently demonstrated associations between hypotension and all-cause mortality in specific subgroups, including age (≥60 years), sex (man), race/ethnicity (non-Hispanic white or non-Hispanic black), diabetes mellitus (yes or no), hypercholesterolemia (yes or no), BMI (≥30kg/m^2^), and eGFR (<90 mL/min/1.73m^2^) subgroups (Figure 3A). However, significant interactions were only observed in the elderly subgroup (age ≥60 years) and the obese subgroup (BMI ≥30kg/m^2^). In terms of CVD mortality, similar results were observed; however, no significant interactions were found in any of the subgroups analyzed (Figure 2B).

**Figure 3.**
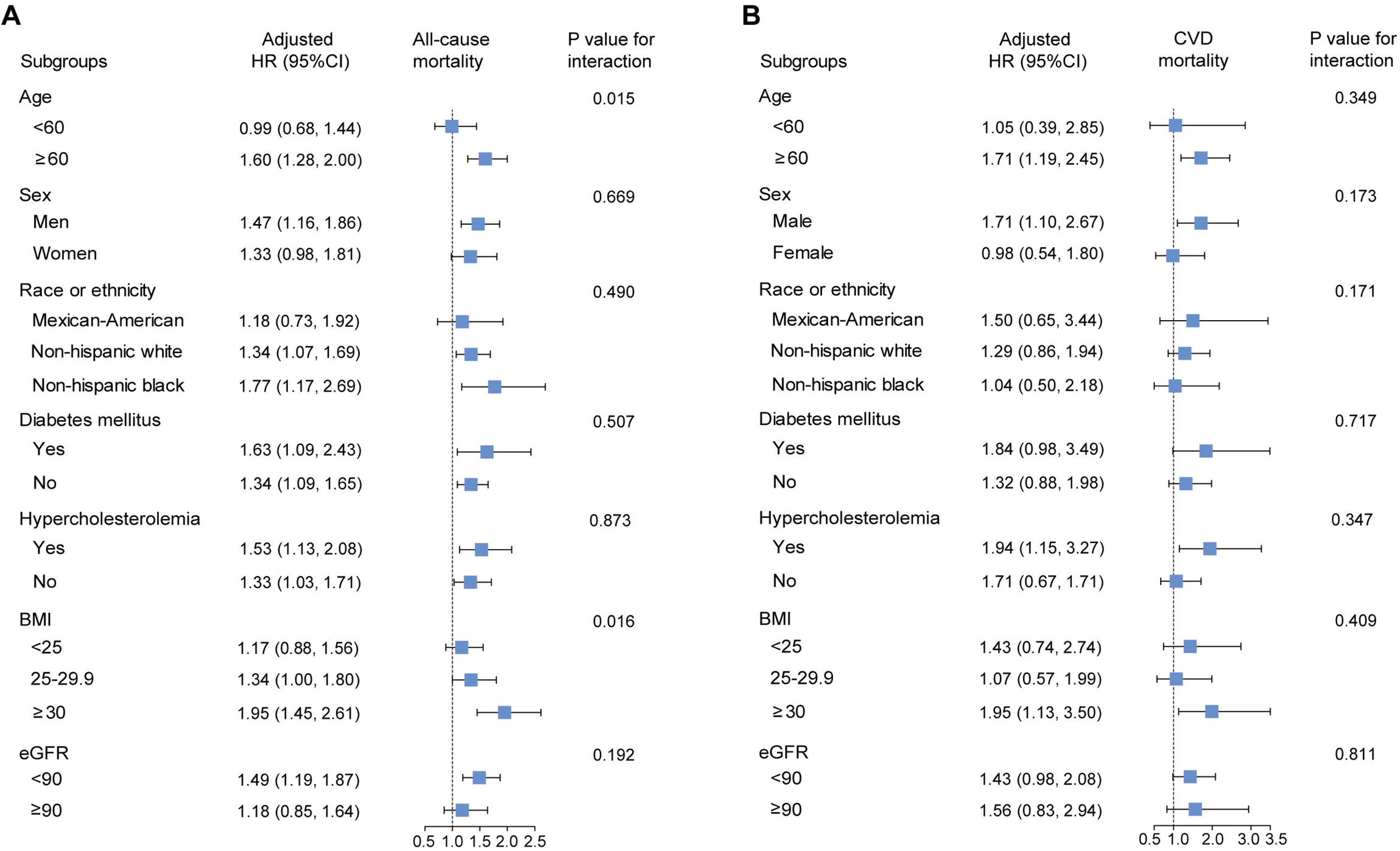
Subgroup analyses of association between hypotension with all-cause and CVD mortality. **(A)** all-cause mortality. **(B)** CVD mortality. HRs (blue box) and 95% CIs (bars on both sides of box) were estimated after adjusting for age, sex, race/ethnicity, education level, marital status, smoking status, drinking status, physical activity, history of CVD, history of chronic lung diseases, history of cancer, diabetes mellitus, hypercholesterolemia, BMI, alanine aminotransferase, and eGFR. Abbreviations: BMI, body mass index; CI, confidence interval; CVD, cardiovascular disease; eGFR, estimated glomerular filtration rate; HR, hazard ratio.

### Sensitivity Analyses

Sensitivity analyses, incorporating the AAC guidelines in 2017 for defining hypertension (eTable 3 in Supplement 1), additionally adjusting for the poverty income ratio and healthy eating index (eTable 4 in Supplement 1), and accounting for deaths during early follow-up years (eTable 5 in Supplement 1), demonstrated that individuals with hypotension exhibited a higher risk of mortality compared to those with normal BP.

## Discussion

In this nationally representative cohort of US adults, hypotension was significantly associated with increased all-cause and CVD mortality, particularly among individuals aged 60 years or older or those with obesity. These findings are of significant importance as they serve to rekindle public awareness regarding this previously overlooked issue.

Numerous pieces of evidence have demonstrated the association between hypertension and all-cause as well as CVD mortality in the general population,^16,18^ which is consistent with our findings. However, there is limited and controversial research on hypotension in the general population.^6–9,19^ Previous studies have observed a U-shaped or J-shaped curve between consecutive blood pressure levels and mortality risk but often attributed this relationship to underlying comorbidities.^8^

In our study, we adjusted for demographic and lifestyle factors and found a significant association between hypotension and mortality risk, aligning with previous researches.^8,19^ Importantly, even after further adjustment for underlying diseases such as CVD, chronic lung diseases, cancer, diabetes mellitus, hypercholesterolemia, and abnormalities in eGFR, ALT, and BMI; the observed association remained significant. This suggests that the increased mortality risk associated with hypotension may not solely be attributed to underlying diseases.

Hypotension is often overlooked due to its asymptomatic nature, but it becomes a concern when inadequate pumping pressure leads to insufficient oxygenated blood perfusion of vital organs. This has been observed in certain patient populations (such as sepsis, critically ill patients, and those undergoing hemodialysis), where hypotension increases the risk of mortality.^10–13^ In our subgroup analyses, we found hypotension interacts with age and BMI, with higher mortality risks observed in elderly and obese individuals. Possible explanations for this result include decreased total cardiac output of the heart, reduced vascular elasticity caused by arteriosclerosis, and overmedication. ^20–22^ Given the aging population and growing obesity problem worldwide,^23,24^ the hypotension, especially in these special populations, should not be neglected any more.

The major strengths of this study lie in the utilization of a large sample size from a nationwide representative cohort in the US. The inclusion of such a substantial sample size not only enhances the persuasiveness of our results but also enables us to conduct subgroup analyses with sufficient statistical power, thereby identifying two high-risk populations for all-cause mortality associated with hypotension: elderly individuals and those with obesity. Furthermore, we have meticulously adjusted for underlying diseases when assessing the mortality risk of hypotension, thus minimizing any potential influence these conditions may have on our findings. Additionally, we have performed a series of sensitivity analyses to demonstrate the robustness and reliability of our conclusions.

This study also has several limitations. First, although we utilized the average BP of three BP measurements to represent the true BP of each participant, it may still introduce measure bias. Ambulatory blood pressure monitoring is recommended to mitigate this bias effectively.^25^ Second, due to the cross-sectional design employed in this study, it cannot provide insights into longitudinal changes in BP over time. Third, self-reported questionnaires were employed for evaluating lifestyle and underlying diseases which may introduce recall bias. Fourth, death certificates may not accurately reflect the precise cause of death. Therefore, a well-designed prospective study conducted in diverse regions would be highly appreciated to validate our findings.

In conclusion, based on a nationally representative sample of US adults spanning from 1999 to 2018, hypotension exhibited significant associations with all-cause and cardiovascular disease mortality, particularly among older individuals and those with obesity. While further validation in independent populations is warranted, these findings underscore the imperative of directing increased attention towards hypotension in the general population. Meanwhile, it is crucial to explore strategies for optimizing the management of hypotension and reducing mortality risk, including lifestyle modifications, drug adjustments, and individualized treatments.

## Data Availability

All data produced are available online at https://www.cdc.gov/nchs/nhanes/index.htm

## Contributors

HR, PH, and ZC designed the study. YH and JT conducted the data analysis. YH and JT drafted the manuscript. NW, ZL, DC, HR, PH, and ZC critically revised the manuscript for important intellectual content. All authors approved the final version of the manuscript. The corresponding author attests that all the listed authors meet authorship criteria and that no others meeting the criteria have been omitted.

## Data sharing

NHANES data are available at www.cdc.gov/nchs/nhis/index.htm.

## Acknowledgments

This work was supported by the Joint Project of Pinnacle Disciplinary Group of The Second Affiliated Hospital of Chongqing Medical University, the Remarkable Innovation-Clinical Research Project of the Second Affiliated Hospital of Chongqing Medical University, the first batch of key disciplines on public health in Chongqing, Health Commission of Chongqing, China, and the Science and technology research project of Chongqing Education Commission (KJZD-K202300404). We gratefully acknowledge the contribution of the participants of the NHANES cohort, research assistants, and facilitating personnel.

