## Supplement 1 for "Association of hypotension with mortality among US adults: prospective cohort study": Supplement 1.docx

**eMethods**

**The definitions of covariates**

Smoking status was categorized as smoker and never smoker. Smoker replied “yes”, and never smoker replied “no” to the question: “Have you smoked at least 100 cigarettes in life?”

Drinking status was categorized as drinker and never drinker. Drinker replied “yes”, and never drinker replied “no” to the question: “Have you ever had at least 12 alcohol drinks in a year?”

Sedentary behavior was defined as if individuals answered ’no’ to all questions about engaging in any of the following physical activities over the last month: vigorous activity in work or recreational time, moderate activity in work or recreational time, tasks around home/yard, and walked or bicycled.

History of cardiovascular disease was defined as a self-reported history of congestive heart failure, coronary heart disease, angina/angina pectoris, heart attack, or stroke.

History of chronic lung disease was defined as a self-reported history of emphysema or chronic bronchitis.

History of cancer was defined as a self-reported history of any cancer.

Diabetes mellitus was defined as fasting glucose ≥126 mg/dl, hemoglobin A1c ≥6.5%, and/or self-reported doctor diagnosis, and/or drug use ^1^.

Hypercholesterolemia was defined as self-reported doctor diagnosis, drug use, or total cholesterol concentration >6.2 mmol/L.^2^

Estimated glomerular filtration rate (eGFR) was calculated using the CKD Epidemiology Collaboration Creatinine Equation, which higher than 90 mL/min/1.73m^2^ was considering normal.^3^

| **Characteristic** | **All participants ^#^**  **(n=55,081)** | **Final enrolled sample**  **(n=37,832)** | **P value** |
| --- | --- | --- | --- |
| Age, years | 47.04 (17.02) | 47.58 (16.88) | <0.001 |
| Sex |  |  | <0.001 |
| Male | 26,473 (48.0%) | 19,070 (49.6%) |  |
| Female | 28,608 (52.0%) | 18,762 (50.4%) |  |
| Race and ethnicity |  |  | <0.001 |
| Mexican American | 9,593 (8.2%) | 6,492 (8.2%) |  |
| Other Hispanic | 4,522 (5.6%) | 3,279 (5.4%) |  |
| Non-Hispanic White | 24,304 (68.0%) | 16,998 (69.6%) |  |
| Non-Hispanic Black | 11,516 (11.3%) | 7,610 (10.2%) |  |
| Other race | 5,146 (7.0%) | 3,453 (6.6%) |  |
| Education level |  |  | <0.001 |
| High school or less | 27,873 (41.7%) | 18,583 (40.3%) |  |
| College or associates degree | 15,366 (30.7%) | 10,968 (31.4%) |  |
| College graduate or above | 11,722 (27.6%) | 8,281 (28.3%) |  |
| Marital status |  |  | 0.005 |
| Married | 32,453 (63.4%) | 22,729 (63.9%) |  |
| Unmarried | 22,038 (36.6%) | 15,103 (36.1%) |  |
| Systolic blood pressure | 122.70 (17.97) | 122.76 (17.53) | 0.001 |
| Diastolic blood pressure | 70.92 (12.46) | 70.96 (12.21) | >0.9 |

Note: Data are shown as the mean (SD) or unweighted frequency counts (weighted percentage) as appropriate. ^#^, all participants aged ≥20 year old in the National Health and Nutrition Examination Survey (NHANES), 1999-2018.

**eTable 2. Associations of all-cause and CVD mortality with each 10 mmHg increase of blood pressure.**

|  | **Hazard ratio (95% CI)** | | | | |
| --- | --- | --- | --- | --- | --- |
|  | **Systolic blood pressure** | |  | **Diastolic blood pressure** | |
| **All-cause mortality** | **<113 mmHg (N=10,369)** | **≥113 mmHg (N=27,463)** |  | **<77 mmHg (N=27,114)** | **≥77 mmHg (N=10,718)** |
| Number of events | 744 | 4517 |  | 4037 | 1224 |
| Crude model | 0.78 (0.65-0.93) | 1.36 (1.34-1.39) |  | 0.73 (0.71-0.75) | 1.28 (1.16-1.42) |
| Model 1 | 0.77 (0.66-0.90) | 1.07 (1.05-1.10) |  | 0.93 (0.90-0.96) | 1.17 (1.06-1.31) |
| Model 2 | 0.79 (0.67-0.94) | 1.08 (1.06-1.10) |  | 0.95 (0.92-0.98) | 1.17 (1.05-1.30) |
| **CVD mortality** | **<112 mmHg (N=9,197)** | **≥112 mmHg (N=28,635)** |  | **<75 mmHg (N=24,557)** | **≥75 mmHg (N=13,275)** |
| Number of events | 171 | 1493 |  | 1219 | 445 |
| Crude model | 0.68 (0.53-0.89) | 1.42 (1.38-1.46) |  | 0.69 (0.66-0.72) | 1.40 (1.23-1.60) |
| Model 1 | 0.72 (0.57-0.91) | 1.1 (1.06-1.13) |  | 0.91 (0.86-0.95) | 1.29 (1.12-1.48) |
| Model 2 | 0.79 (0.61-1.03) | 1.11 (1.07-1.14) |  | 0.93 (0.88-0.97) | 1.28 (1.11-1.48) |

Abbreviations: CI, confidence interval; CVD, cardiovascular disease. Crude Model, no covariates were adjusted. Model 1 was adjusted for age, sex, race and ethnicity, education level, marital status, smoking, drinking, and physical activity. Model 2 was further adjusted for history of cardiovascular diseases, history of chronic lung disease, history of cancer, diabetes mellitus, hypercholesterolemia, body mass index, estimated glomerular filtration rate, and alanine aminotransferase in addition to Model 1.

**eTable 3. Associations of all-cause and CVD mortality with different classifications of BP refereeing to ACC guideline ^a^.**

|  | **Hazard ratio (95% CI)** | | | |
| --- | --- | --- | --- | --- |
|  | **Hypotension (N=2,856)** | **Normal BP (N=16,978)** | **High Normal BP (N=3,536)** | **Hypertension (N=14,462)** |
| **All-cause mortality** | | | | |
| Number of events | 259 | 437 | 449 | 4116 |
| Crude model | 2.06 (1.69-2.51) | 1 (reference) | 2.15 (1.83-2.54) | 5.40 (4.81-6.05) |
| Model 1 | 1.59 (1.30-1.95) | 1 (reference) | 1.05 (0.91-1.22) | 1.35 (1.19-1.53) |
| Model 2 | 1.51 (1.23-1.85) | 1 (reference) | 1.22 (1.04-1.42) | 1.32 (1.16-1.50) |
| **CVD mortality** | | | | |
| Number of events | 70 | 93 | 115 | 1386 |
| Crude model | 2.77 (1.92-4.00) | 1 (reference) | 2.65 (1.97-3.58) | 9.22 (7.17-11.9) |
| Model 1 | 1.86 (1.28-2.72) | 1 (reference) | 1.26 (0.96-1.67) | 1.85 (1.42-2.40) |
| Model 2 | 1.74 (1.18-2.56) | 1 (reference) | 1.29 (0.96-1.73) | 1.68 (1.28-2.21) |

Abbreviations: ACC, American College of Cardiology; BP, blood pressure; CI, confidence interval; CVD, cardiovascular disease. Crude Model, no covariates were adjusted. Model 1 was adjusted for age, sex, race and ethnicity, education level, marital status, smoking, drinking, and physical activity. Model 2 was further adjusted for history of cardiovascular diseases, history of chronic lung disease, history of cancer, diabetes mellitus, hypercholesterolemia, body mass index, estimated glomerular filtration rate, and alanine aminotransferase in addition to Model 1. ^a^ According to ACC hypertension guideline in 2017, hypertension is defined as SBP ≥130 mmHg or DBP ≥80 mmHg. The high normal BP is defined as 120-129 and <80 mmHg, which companied with the change of normal BP, i.e., 90-119 and 60-79 mmHg.

**eTable 4. Associations of all-cause and CVD mortality with different classifications of BP in models further adjusted for covariates of PIR and HEI ^a^.**

|  | **Hazard ratio (95% CI)** | | | |
| --- | --- | --- | --- | --- |
|  | **Hypotension (N=2,542)** | **Normal BP (N=15,289)** | **High Normal BP (N=3,167)** | **Hypertension (N=12,827)** |
| **All-cause mortality** | | | | |
| Number of events | 222 | 841 | 413 | 3205 |
| Crude model | 1.60 (1.34-1.91) | 1 (reference) | 2.29 (1.93-2.72) | 5.69 (5.20-6.23) |
| Model 1 | 1.54 (1.28-1.84) | 1 (reference) | 1.05 (0.89-1.24) | 1.40 (1.27-1.55) |
| Model 2 | 1.45 (1.20-1.75) | 1 (reference) | 1.10 (0.93-1.30) | 1.33 (1.20-1.47) |
| Model 3 | 1.43 (1.18-1.73) | 1 (reference) | 1.10 (0.93-1.31) | 1.32 (1.20-1.47) |
| **CVD mortality** | | | | |
| Number of events | 60 | 199 | 127 | 1105 |
| Crude model | 1.91 (1.33-2.74) | 1 (reference) | 3.01 (2.23-4.06) | 8.96 (7.49-10.7) |
| Model 1 | 1.70 (1.17-2.46) | 1 (reference) | 1.23 (0.92-1.64) | 1.84 (1.49-2.26) |
| Model 2 | 1.57 (1.07-2.30) | 1 (reference) | 1.28 (0.96-1.70) | 1.58 (1.28-1.96) |
| Model 3 | 1.57 (1.07-2.29) | 1 (reference) | 1.29 (0.96-1.72) | 1.58 (1.28-1.96) |

Abbreviations: BP, blood pressure; CI, confidence interval; CVD, cardiovascular disease; HEI, healthy eating index; PIR, poverty income ratio. Crude Model, no covariates were adjusted. Model 1 was adjusted for age, sex, race and ethnicity, education level, marital status, smoking, drinking, and physical activity. Model 2 was further adjusted for history of cardiovascular diseases, history of chronic lung disease, history of cancer, diabetes mellitus, hypercholesterolemia, body mass index, estimated glomerular filtration rate, and alanine aminotransferase in addition to Model 1. Model 3 was further adjusted for PIR and HEI in addition to Model 2. ^a^ We further excluded participants without data on PIR (n=3 108) and HEI (n=899) from the entire participants, and 33 825 individuals were included in this sensitivity analysis.

**eTable 5. Associations of all-cause and CVD mortality with different classifications of BP after excluding death within the first years of follow-up.**

|  | **Hazard ratio (95% CI)** | | | |
| --- | --- | --- | --- | --- |
|  | **Hypotension (N=2,833)** | **Normal BP (N=16,927)** | **High Normal BP (N=3,515)** | **Hypertension (N=14,244)** |
| **All-cause mortality** | | | | |
| Number of events | 236 | 897 | 440 | 3375 |
| Crude model | 1.62 (1.34-1.96) | 1 (reference) | 2.35 (2.01-2.76) | 5.68 (5.22-6.19) |
| Model 1 | 1.54 (1.28-1.86) | 1 (reference) | 1.07 (0.92-1.24) | 1.38 (1.26-1.51) |
| Model 2 | 1.44 (1.19-1.76) | 1 (reference) | 1.11 (0.95-1.30) | 1.30 (1.19-1.43) |
| **CVD mortality** | | | | |
| Number of events | 63 | 207 | 134 | 1164 |
| Crude model | 1.96 (1.38-2.79) | 1 (reference) | 3.15 (2.36-4.20) | 9.02 (7.56-10.8) |
| Model 1 | 1.71 (1.18-2.47) | 1 (reference) | 1.26 (0.96-1.66) | 1.80 (1.46-2.21) |
| Model 2 | 1.57 (1.07-2.29) | 1 (reference) | 1.31 (1.00-1.73) | 1.55 (1.26-1.91) |

Abbreviations: BP, blood pressure; CI, confidence interval; CVD, cardiovascular disease. Crude Model, no covariates were adjusted. Model 1 was adjusted for age, sex, race and ethnicity, education level, marital status, smoking, drinking, and physical activity. Model 2 was further adjusted for history of cardiovascular diseases, history of chronic lung disease, history of cancer, diabetes mellitus, hypercholesterolemia, body mass index, estimated glomerular filtration rate, and alanine aminotransferase in addition to Model 1.

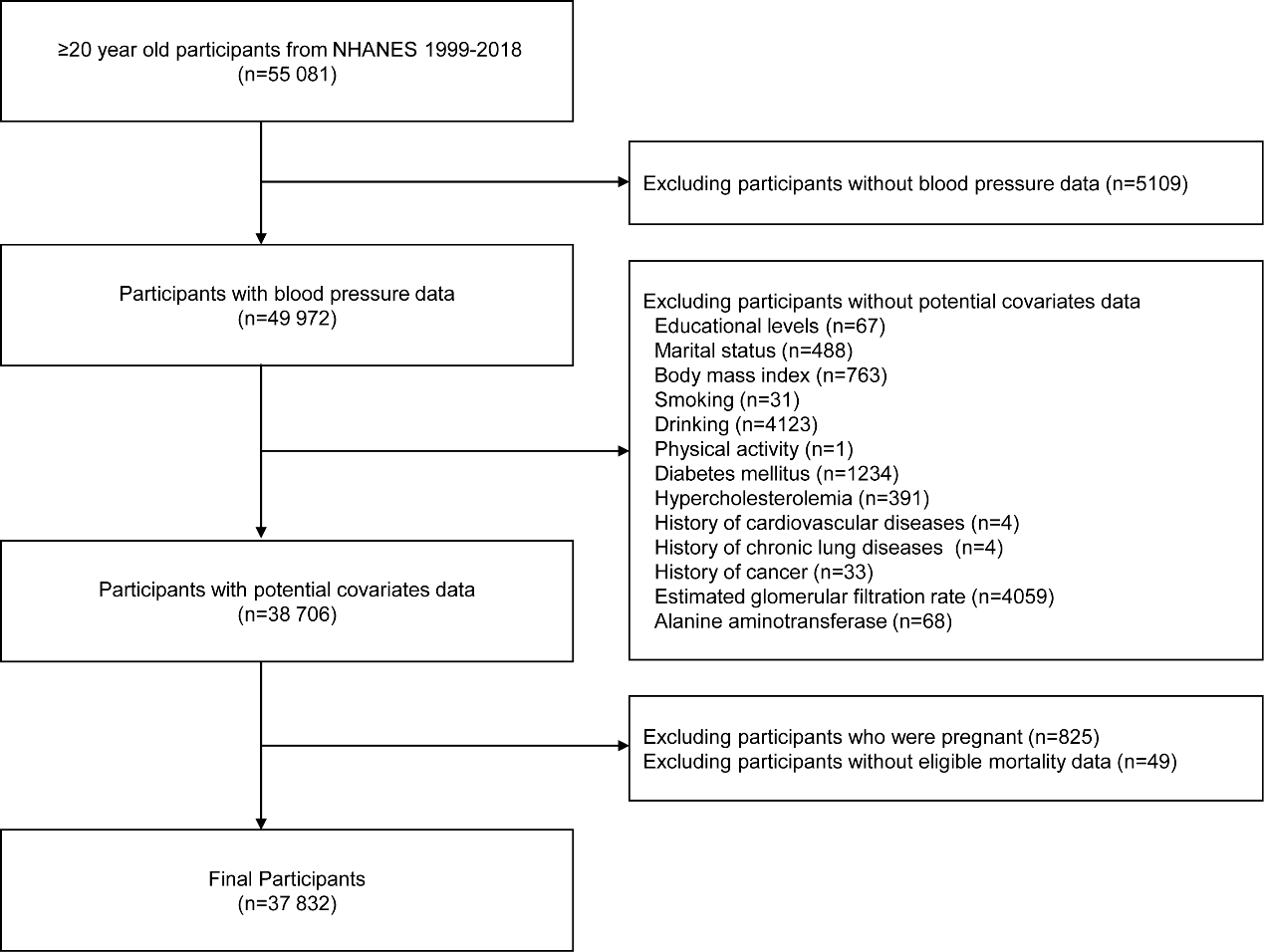

**eFigure 1. Selection of study participants from the National Health and Nutrition Examination Survey, 1999 to 2018.**
